# Lung-Kidney Axis in Cystic Fibrosis: Early Urinary Markers of Kidney Injury Correlate with Neutrophil Activation and Worse Lung Function

**DOI:** 10.1101/2023.11.10.23298378

**Authors:** Grace M. Rosner, Himanshu B. Goswami, Katherine Sessions, Lindsay K. Mendyka, Brenna Kerin, Irma Vlasac, Diane Mellinger, Lorraine Gwilt, Thomas H. Hampton, Martha Graber, Alix Ashare, William T. Harris, Brock Christensen, Bruce A. Stanton, Agnieszka Swiatecka-Urban, Sladjana Skopelja-Gardner

## Abstract

**Background:** Adult people with cystic fibrosis (PwCF) have a higher risk of end-stage kidney disease than the general population. The nature and mechanism of kidney disease in CF are unknown. This study quantifies urinary kidney injury markers and examines the hypothesis that neutrophil activation and lung infection are associated with early kidney injury in CF.

**Methods:** Urinary total protein, albumin, and markers of kidney injury and neutrophil activation, normalized to creatinine, as well as urinary immune cells, were quantified in CF (n = 48) and healthy (n = 33) cohorts. Infection burden and chronicity were defined by sputum culture and serum titers of anti-bacterial antibodies.

**Results:** PwCF had increased urinary protein levels, consisting of low-molecular-weight tubular injury markers, independent of glomerular filtration rate (eGFR). This finding suggests subclinical renal injury processes. Urinary analysis of the CF cohort identified different associations of urinary injury markers with aminoglycoside exposure, lung function, and neutrophil activation. High urinary KIM-1 levels and increased prevalence of neutrophils among urine immune cells correlated with decreased lung function in PwCF. The relationship between tubular injury and decreased lung function was most prominent in patients harboring chronic *Pseudomonas aeruginosa* infection.

**Conclusions:** Increased urinary tubular injury markers in PwCF suggest early subclinical renal injury not readily detected by eGFR. The strong association of high urinary KIM-1 and neutrophils with diminished lung function and high *Pseudomonas aeruginosa* burden suggests that pulmonary disease may contribute to renal injury in CF.

## Introduction

Even though lung disease is the primary cause of morbidity and mortality in CF, extrapulmonary manifestations in the pancreas, intestine, or kidney may contribute to disease outcomes. Advancements in early diagnosis, antibiotic treatment, and most notably, high-efficiency CFTR modulator treatments (HEMT) have remarkably extended the lifespan of people with CF (PwCF). This increased longevity has brought to light a rise in long-term complications, such as chronic kidney disease (CKD), which affects 1 in 7 adults in the United States. The literature estimates that up to 29% of PwCF suffer from CKD ^1–3^. The risk of CKD doubles every 10 years of follow-up in PwCF, and PwCF with CKD have a higher risk of developing end-stage renal disease (ESRD) compared to non-CF patients with CKD ^2,4^. Despite these alarming figures, the nature of early renal injury and the mechanisms by which inflammatory events in the lung may contribute to kidney pathogenesis in CF remain unknown.

The two most studied contributors to kidney disease in CF are aminoglycosides, used to treat pulmonary exacerbations triggered by *Pseudomonas aeruginosa* (*P. aeruginosa*) infections, and CF-related diabetes (CFRD) ^2,5^. Although diabetes is a significant risk of CKD in both non-CF and CF populations ^2^, it is a less frequent cause of ESRD in CF (24% in CF populations vs. 42% in non-CF populations, p<0.001) ^4^. Aminoglycosides disrupt tubular reabsorption, leading to tubular necrosis and acute kidney injury (AKI) ^6^, though several studies showed that kidney function in PwCF may be independent of prolonged aminoglycoside use ^2,7,8^. How infection and lung inflammation may contribute to CKD in PwCF has not been extensively studied. That inflammatory events in the lung can cause kidney injury was first recognized in acute respiratory disease syndrome (ARDS)-mediated AKI, but likely extends to other lung illnesses ^9^. In chronic obstructive pulmonary disease (COPD), ∼24% of patients had persistent albuminuria, compared to 4% of control subjects, and lower estimated glomerular filtration rate (eGFR) in COPD patients correlated with decreased lung function ^10^. In *in vivo* murine studies, intratracheal instillation of lipopolysaccharide (LPS) caused acute renal injury accompanied by both pulmonary and renal inflammation ^11^. Whether infection-triggered inflammation or diminished lung function in CF contribute to early kidney injury is unknown. Given the high neutrophil infiltration in the CF lung before and during infection ^12–14^, and the reported ability of neutrophils to mediate both local and distal tissue damage ^15,16^, we hypothesized that activated neutrophils contribute to kidney injury in PwCF.

Several studies have reported increased levels of a limited number of urinary kidney injury markers in PwCF ^3,17–19^, suggesting that kidney damage may develop before it is detected by standard clinical tests, e.g. albuminuria or eGFR. Neither a decrease in eGFR, which may occur as late as after ≥50% of kidney function loss, nor an increase in albuminuria, a marker of glomerular dysfunction, provide an insight into disease etiology or capture early signs of renal tubular injury. This study aims to define early urinary markers of kidney damage and asks if lung infection and neutrophil-mediated inflammation may play a role in subclinical renal injury in CF.

## Methods

### Study Cohorts

Urine and blood were collected by written informed consent from a cohort of 48 adult (> 18 years old, Cohort 1) PwCF, recruited between the years 2017 and 2022, at Dartmouth-Hitchcock Medical Center (DHMC), Lebanon, New Hampshire, through the Clinical and Translational Research Core (CTRC) of the Dartmouth CF P30. Frozen urine and serum samples and demographic and clinical data were obtained from the CTRC. Sputum cultures, patient lung function, and other laboratory measurements were assayed by clinical laboratories at DHMC. eGFR was computed using the CKD-EPI Creatinine Equation. Urine from 33 healthy controls (HC) was collected by written informed consent. Cohort characteristics are described in Supplemental Tables 1 and 2. An additional CF cohort of 11 patients (Cohort 2: 2022-2023, characteristics in Supplemental Table 3) was recruited to obtain urinary cell pellets for methylation studies.

### Urine collection and processing

Urine samples were processed immediately after collection and tested for urinary tract infections using Chemstrip^®^ Test Strips (Roche). Samples used for urinary analysis of kidney injury and neutrophil activation were aliquoted and immediately frozen at −80°C. For urine DNA methylation studies, urine samples were centrifuged (200*g*) to separate cell-free urine from the cell pellet. Pellets were preserved at −80°C.

### Quantification of urinary renal injury markers and neutrophil activation

Total urine protein levels were quantified with Bradford assay (Pierce™). Urinary albumin and creatinine levels were determined using the Albuwell Hu and Creatinine Companion ELISA kit (Ethos Biosciences). Levels of urinary kidney injury proteins were quantified by ELISA: VCAM-1 (R&D); podocalyxin (Invitrogen); beta-NAG (Abcam); or Human Kidney Biomarker Panels Luminex® (R&D): Panel 1: KIM-1, Cystatin-C, NGAL; Panel 2: β2MG, EGF, TFF-3, Osteopontin. S100A8/A9 was measured by ELISA (R&D). NETs were quantified by ELISA measuring MPO: DNA complexes ^20^. All protein concentrations were normalized to urinary creatinine.

### Anti-*P. aeruginosa* and anti-*S. aureus* immunoglobulin G (IgG) quantification

Serum concentrations of anti-*P. aeruginosa* and anti-*Staphylococcus aureus (S. aureus)* IgG were quantified by ELISA using bacterial lysates generated from overnight cultures of *P. aeruginosa* PA14 and *S. aureus* USA100 strains ^21^.

### DNA methylation (DNAm)-based deconvolution of cell types in urine

DNA from urine cell pellets was extracted using Zymogen Mini/Micro Prep Kit and 250 ng bisulfide converted and run on the EPIC Illumina DNAm array (Dartmouth Genomics Core). Infinium Methylation EPIC BeadChip v1.0 (EPIC) raw intensity data (IDAT) files were preprocessed using minfi and Enmix. Probes with out-of-band hybridization >0.05 were excluded, followed by normalization and background correction using *FunNorm, (*minfi). Probe type correction for EPIC was completed using BMIQ. EpiDISH 2.12/HEpiDISH was used to calculate cell type proportions ^22^. We used an immune-specific reference data set based on the iterative algorithm Identifying Optimal Libraries (IDOL) ^22^. Robust partial correlation methods were used. In Supplemental Figure 1, we demonstrate that neutrophils cluster together based on the DNAm signature regardless of the biospecimen (blood or urine).

### Statistical analysis

All analyses were performed using GraphPad Prism software and R version 2023.06.0+421. Differences between two means were analyzed using Student’s t-test with a statistical significance of p<0.05. Shapiro-Wilk test was performed to determine the normality of the data. Differences between the two means were analyzed using Student’s t-test for normally distributed data or a non-parametric t-test for not normally distributed data, with a statistical significance of p<0.05. Correlations were assessed using Pearson’s (normally distributed data) or non-parametric Spearman’s analysis (ranked or not-normally distributed data) or Linear Regression analysis to control for potential confounders. Robust Principal component analysis ^23^, to account for not normally distributed data, was performed using urine proteomic values as well as clinical factors.

## Results

### Proteinuria and urinary tubular injury markers are elevated in PwCF

Adult PwCF (n=48) and healthy controls (HC; n=33) were recruited for this study (**Supplemental Tables 1-2**). Total urinary protein, normalized to creatinine, was significantly higher in 29.2% (N = 14) of PwCF, compared to HC (**Fig. 1A**, **Table 1**). Urinary protein did not correlate with eGFR (**Fig. 1A**). The increased urinary protein in PwCF was not due to albuminuria, as the urine albumin/creatinine ratio was comparable between the CF and HC cohorts (**Fig. 1B**).

**Figure 1.**
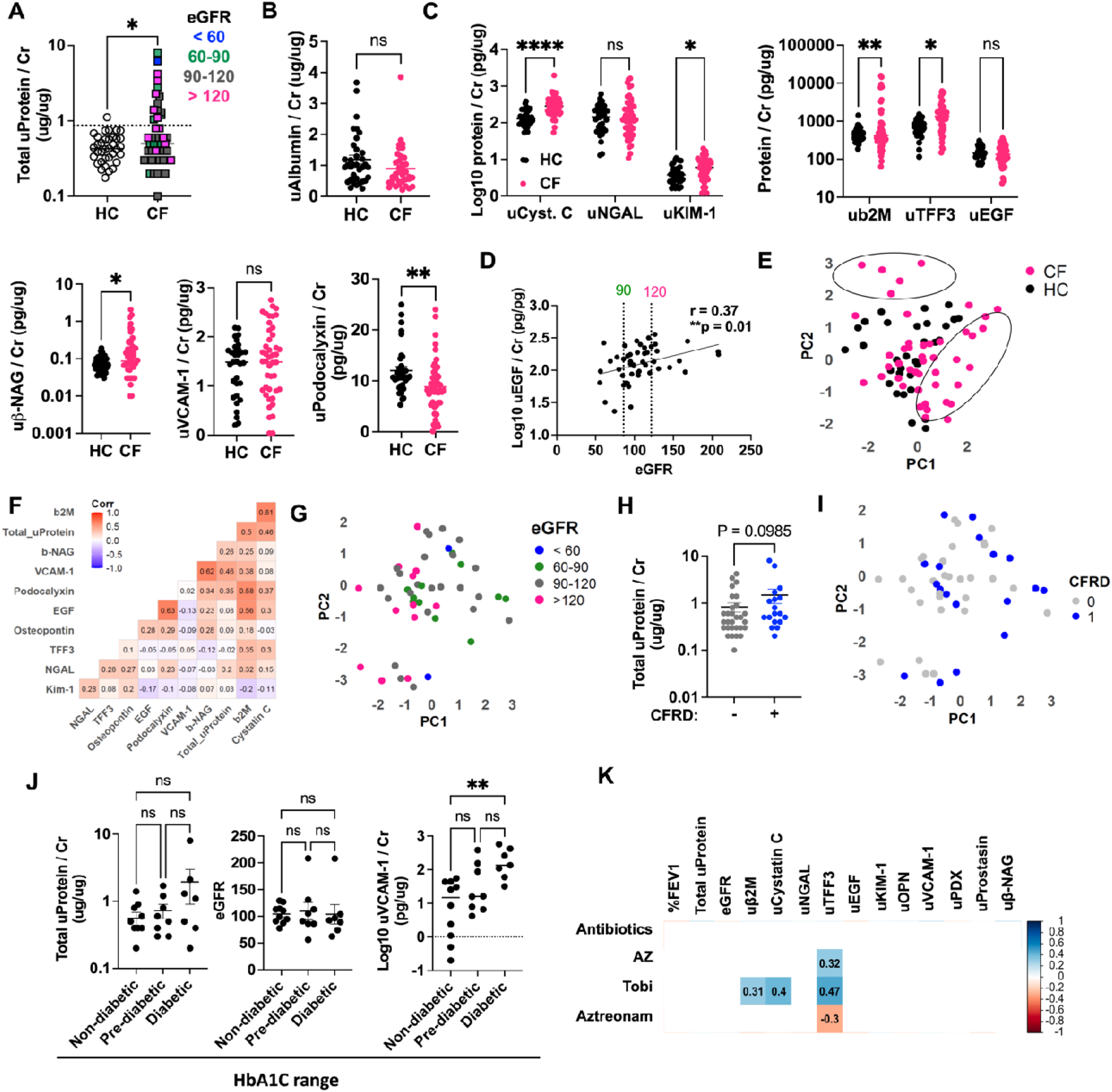
Urinary levels of tubular proteins indicate subclinical renal injury in adult PwCF. **(A)** Total urine protein levels were measured by Bradford assay and normalized to urine creatinine in health control (HC, n =33) and CF (CF, n= 48) cohorts and stratified by eGFR, computed using the CKD-EPI Creatinine Equation: blue: <60, green: 60-90, grey: 90-120, pink: >120. Significance was defined by a non-parametric t-test, **p < 0.01. The dotted line represents a positive cutoff (0.87) based on mean HC (0.45) ± 2 S.D. (0.21). **(B)** Urinary levels of albumin (uAlbumin) normalized to creatinine in HC and CF cohorts. Non-parametric-test defined significance, ns = not significant (p > 0.05). **(C)** Urinary levels of kidney injury markers in HC (pink) and CF (black) cohorts, normalized to urine creatinine. Significance was determined by mixed effects analysis multiple comparisons with Bonferroni correction (Luminex panel 1: Cystatin C, NGAL, KIM-1; Luminex panel 2: b2M, TFF3, EGF); non-parametric t-test (bNAG, PDX); student’s t-test (VCAM1) **(D)** Pearson correlation analysis between urinary levels of epidermal growth factor (log10 uEGF), normalized to urine creatinine, and eGFR, computed as in A. **(E)** Robust principal component (PC) analysis of urinary kidney injury markers measured in panel C in HC and CF cohorts. Circles indicate two subpopulations of the CF cohort with differential urinary levels of injury markers. **(F)** Spearman correlation analysis of urinary injury markers and total urine protein, normalized to urine creatinine, in the CF cohort. (**G**) Robust PCA of urinary markers measured in panel C in the CF cohort, as a function of eGFR. **(H)** total urine protein normalized to creatinine, in PwCF with (pink) or without (grey) a diagnosis of CF-related diabetes (CFRD), were compared by non-parametric t-test. **(I)** Robust principal component (PC) analysis of urinary markers measured in C in the CF cohort stratified by diagnosis of CFRD (blue) or no diagnosis of CFRD (grey). **(J)** Total urine protein levels, eGFR, and uVCAM-1 levels in PwCF stratified by HbA1c levels: non-diabetic (< 5.7), pre-diabetic (5.7 – 6.5), and diabetic (>6.5); non-parametric t-test (total urine protein) and student’s t-test (eGFR, uVCAM1); ns = not-significant, p > 0.05; **p < 0.01. **(J)** Spearman correlation analysis between urinary kidney injury markers measured in C, % FEV1, eGFR, and status of antibiotic, azithromycin (AZ), tobramycin (Tobi), or aztreonam use at the time of sample collection. The color of the squares indicates a significant positive (blue) or negative (red) correlation, and the r coefficient is shown within each square. β2m = β2-microglobulin, Kim-1 = kidney injury marker 1, EGF = epidermal growth factor, OPN = osteopontin, NGAL = Lipocalin 2, TFF3 = Trefoil factor 3, PDX = Podocalyxin, βNAG = N-acetyl-β-d-glucosaminidase.

**Table 1:**
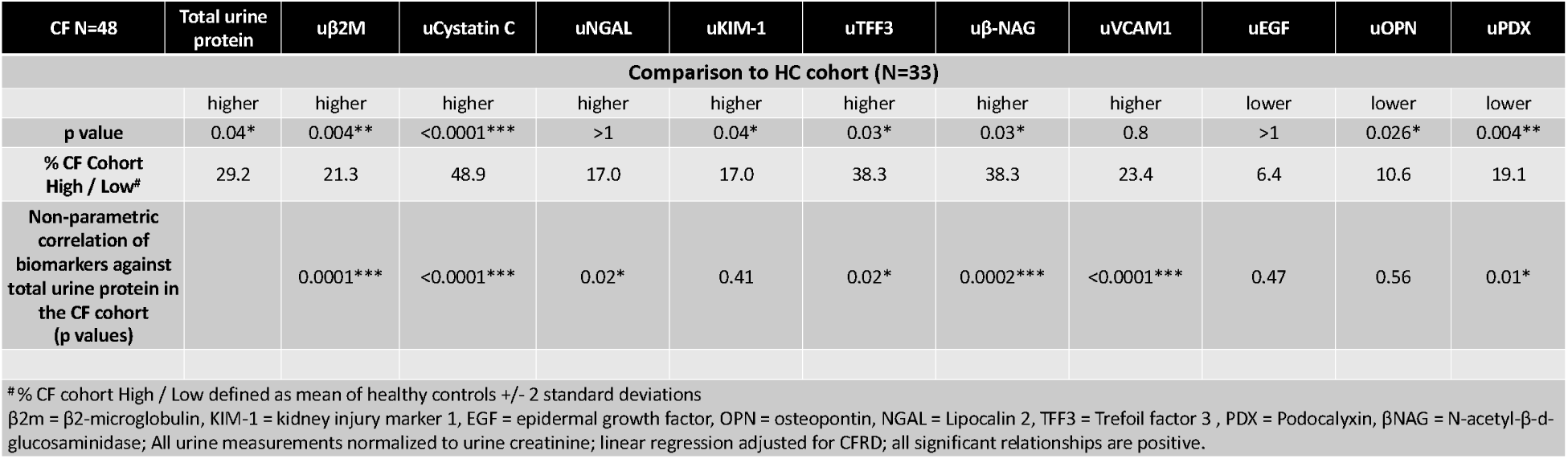
Urinary kidney injury markers in the CF vs. HC cohorts.

| CF N=48 | Total urine protein | u$\beta 2m$ | uCystatin C | uNGAL | uKIM-1 | uTFF3 | uβ-NAG | uVCAM1 | uEGF | uOPN | uPDX |
| --- | --- | --- | --- | --- | --- | --- | --- | --- | --- | --- | --- |
| Comparison to HC cohort (N=33) |  |  |  |  |  |  |  |  |  |  |  |
|  | higher | higher | higher | higher | higher | higher | higher | higher | lower | lower | lower |
| p value | 0.04* | 0.004** | <0.0001*** | >1 | 0.04* | 0.03* | 0.03* | 0.8 | >1 | 0.026* | 0.004** |
| % CF Cohort High / Low* | 29.2 | 21.3 | 48.9 | 17.0 | 17.0 | 38.3 | 38.3 | 23.4 | 6.4 | 10.6 | 19.1 |
| Non-parametric correlation of biomarkers against total urine protein in the CF cohort (p values) |  | 0.0001*** | <0.0001*** | 0.02* | 0.41 | 0.02* | 0.0002*** | <0.0001*** | 0.47 | 0.56 | 0.01* |
| *% CF cohort High / Low defined as mean of healthy controls ± 2 standard deviations<br>$\beta 2m$ = $\beta 2$-microglobulin, KIM-1 = kidney injury marker 1, EGF = epidermal growth factor, OPN = osteopontin, NGAL = Lipocalin 2, TFF3 = Trefoil factor 3, PDX = Podocalyxin, βNAG = N-acetyl-β-d-glucosaminidase; All urine measurements normalized to urine creatinine; linear regression adjusted for CFRD; all significant relationships are positive. | | | | | | | | | | | |

To test if elevated urinary protein levels indicate subclinical tubular injury and to identify early biomarkers of such processes, we used Luminex renal injury panels and ELISA assays to quantify urinary levels of low-molecular-weight kidney injury markers. As summarized in **Table 1**, urinary levels of tubular injury markers, normalized to urine creatinine, including β2MG, Cystatin C, KIM-1, TFF3, and β-NAG, were significantly higher in PwCF compared to HC (**Fig. 1C**). Using the HC to define the baseline urinary levels of each biomarker (mean of HC +/− 2 S.D.), we found that 17–48.9% of CF samples had high urinary levels of one or more kidney injury markers (**Table 1**). Consistent with signs of tubular injury, there was a trend in reduced urinary epidermal growth factor (uEGF) in PwCF (**Table 1**), which positively correlated with eGFR (**Fig. 1D**). Levels of other tubular injury markers did not correlate with eGFR, further supporting that eGFR fails to reflect early signs of tubular damage.

Principal component analysis (PCA) based on urinary measurements of the 10 kidney injury markers demonstrated 33.3% (n=16/48) of PwCF grouped separately from HC (**Fig. 1E**) into two populations (encircled). Correlation analyses showed that most tubular injury markers positively correlated with total urinary protein levels (**Fig. 1F** and **Table 1**) and some correlations between the urinary injury markers were noted: i) β2MG, Cystatin C, VCAM-1, and β-NAG and ii) EGF, PDX, and Cystatin C (**Fig. 1F**). PCA did not reveal distinct clustering of individuals based on the urinary protein concentrations of the kidney injury markers in relation to eGFR (**Fig. 1G**). Moreover, when patients with eGFR < 90, who may already have signs of a declining kidney function, were excluded from analysis, urinary KIM-1, Cystatin C, TFF3, and β-NAG were still significantly higher in the CF compared to the HC cohort (Supplemental Figure 2). These findings suggest a heterogeneity in the urinary profile of PwCF that is not directly correlated with the clinical measure of kidney function, eGFR.

We considered the potential role of CFRD as it is one of the leading causes of CKD in the general population. Despite its high prevalence in PwCF (37.5%, Supplemental Table 1), CFRD diagnosis was not associated with increased total urine protein/creatinine ratio, though a trend in higher proteinuria in CFRD+ patients was noted (Supplemental Table 4 and **Fig. 1G**). PCA of the urinary levels of kidney injury markers did not reveal a distinct separation of individuals based on their CFRD status (**Fig. 1H**). When using serum hemoglobin A1C (HbA1C) levels to stratify PwCF into non-diabetic (< 5.7), pre-diabetic (5.7–6.5), and diabetic (>6.5) groups, we detected significantly increased uVCAM-1 in the diabetic group, but no association was found with total urine protein, eGFR, or other urinary kidney markers (**Fig. 1J**, Supplemental Table 4).

Given the nephrotoxic nature of aminoglycosides, we next asked if increased urinary levels of tubular injury markers in PwCF are a consequence of treatment with tobramycin (nephrotoxic aminoglycoside, 25% of PwCF), compared to antibiotics with low nephrotoxic risk: azithromycin (41.6% of PwCF) and aztreonam (4.1% of PwCF). Urinary levels of β2MG, cystatin C, and TFF3 were significantly increased in PwCF receiving tobramycin therapy at the time of urine collection, while only uTFF3 was elevated in PwCF receiving azithromycin (**Fig. 1K** and Supplemental Table 4). Together, these findings demonstrate early signs of renal injury in PwCF that are partially associated with glucose dysregulation or aminoglycoside therapy but also uncover increased levels of renal injury markers that in this patient cohort are not associated with either of these two factors.

### Decreased lung function and chronic P. aeruginosa infection are associated with markers of early tubular injury in PwCF

While lung-kidney crosstalk has been recognized in several conditions of lung inflammation ^24^, how these organs may communicate in CF patients is unknown. To establish a link between the lung and the kidney in CF we examined if lung function (%FEV_1_) correlates with urinary levels of kidney injury markers. We found that uKIM-1 levels, normalized to creatinine, negatively correlated with %FEV_1_ (**Fig. 2A**), suggesting an inverse relationship between lung function and kidney injury in PwCF. Low %FEV_1_ was most notable in PwCF with a high burden of mucoid *P. aeruginosa* by sputum culture (**Fig. 2A**, red and blue circles).

**Figure 2.**
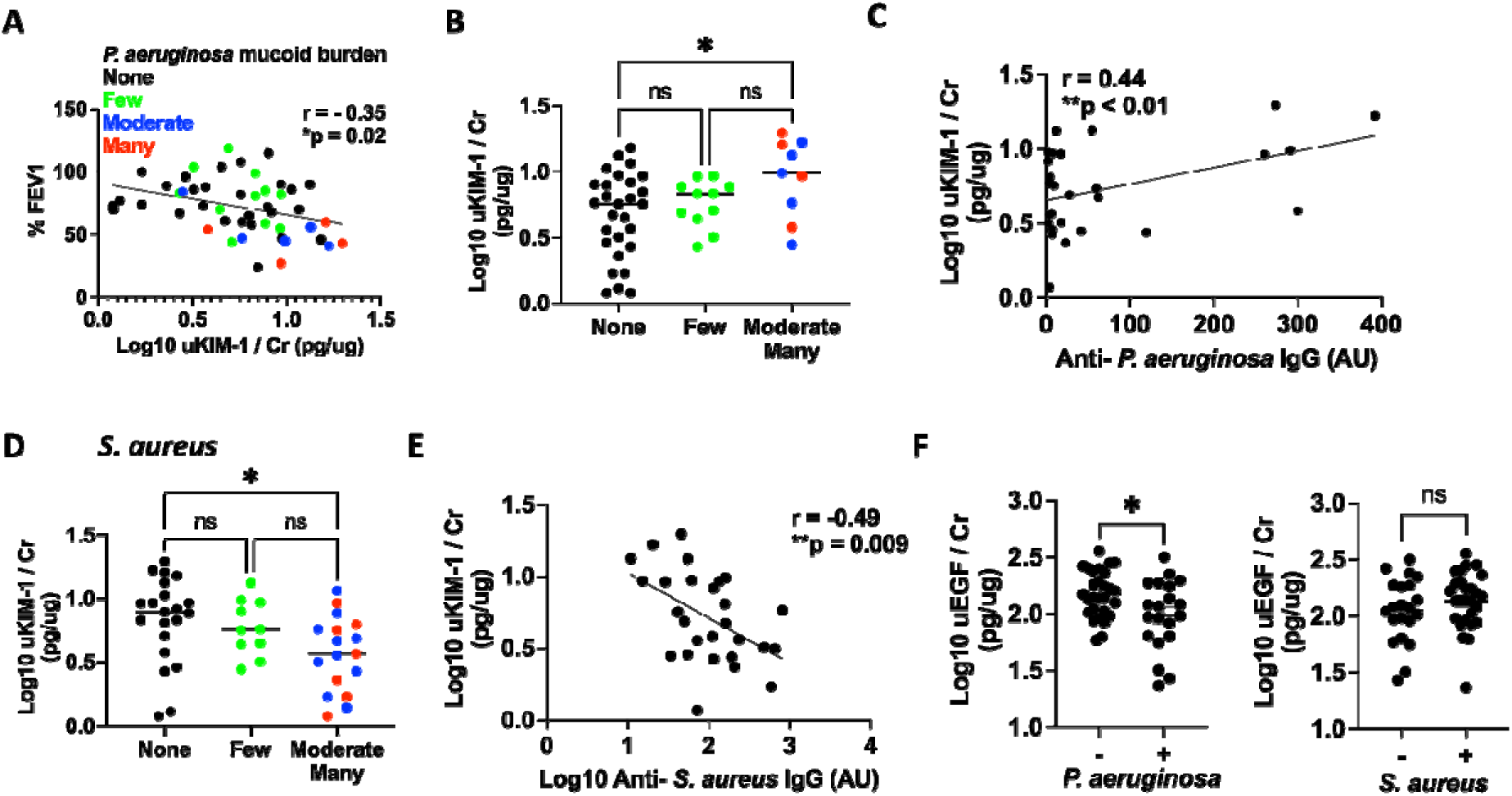
High urinary KIM-1 levels are associated with worse lung function and increased mucoid *P. aeruginosa* burden in PwCF. **(A)** Pearson correlation analysis between urinary KIM-1 (uKIM-1) normalized to urine creatinine and percent predicted forced expiratory volume in 1 second (%FEV1, n =48) in the CF cohort. Distribution of the mucoid *P. aeruginosa* burden (none = black, few = green, moderate = blue, many = red) in sputum culture in relation to %FEV1 and uKIM-1. **(B)** Levels of uKIM-1 in PwCF with no (none), few, and moderate/many mucoid *P. aeruginosa* colonies in the sputum culture were compared by one-way ANOVA with Bonferroni post-hoc (ns = not significant, p > 0.05; *p < 0.05). **(C)** Non-parametric Spearman correlation analysis between uKIM-1 levels and anti-*P. aeruginosa* IgG serum titres (n = 27 PwCF). **(D)** Levels of uKIM-1 in PwCF with no (none), few, and moderate/many *S. aureus* colonies in the sputum culture were compared by one-way ANOVA with Bonferroni post-hoc (ns = not significant, p > 0.05; *p < 0.05). **(E)** Non-parametric Spearman correlation analysis between uKIM-1 levels and anti-*S. aureus* IgG serum titres (n = 27 PwCF). **(F)** Urinary levels of EGF (uEGF), normalized to urine creatinine, in PwCF colonized with *P. aeruginosa* (left) or *S. aureus* (right) b sputum culture were compared to PwCF not colonized with each pathogen by Student’s t-test (n=48, ns = not significant, p > 0.05; *p < 0.05).

Additionally, PwCF with increased sputum burden of mucoid *P. aeruginosa* had significantly higher uKIM-1 than those with lower infection burden (**Fig. 2B**). No association was found between non-mucoid *P. aeruginosa* in sputum and %FEV_1_ or uKIM-1 levels (not shown). In support of a relationship between chronic *P. aeruginosa* infection and renal injury, we found that anti-*P. aeruginosa* IgG levels in the serum of PwCF positively correlated with uKIM-1 levels (**Fig. 2C**). In contrast, PwCF who had higher sputum *S. aureus* burden had significantly lower uKIM-1 compared to PwCF with no or low *S. aureus* burden (**Fig. 2D**), suggesting that chronic *S. aureus* infection may not be associated with kidney injury in CF. Indeed, anti-*S. aureus* IgG levels in the serum were negatively correlated with uKIM-1 levels (**Fig. 2E**). These findings suggest that chronic lung infections with mucoid *P. aeruginosa* strains may be important mediators of the lung-kidney axis in CF. In support of this model, patients with mucoid *P. aeruginosa,* but not *S. aureus,* positive sputum culture also had significantly lower levels of uEGF (**Fig. 2F**). To uncouple the potential contribution of aminoglycoside therapy to higher uKIM-1 and lower uEGF in PwCF infected with *P. aeruginosa*, we asked if these differences were most prominent in patients treated with tobramycin. In our cohort, infections among patients treated with tobramycin were evenly divided between *P. aeruginosa* and *S. aureus.* Tobramycin-treated patients did not preferentially demonstrate higher uKIM-1 or lower uEGF levels (Supplemental Figure 3). Furthermore, linear regression analysis adjusted for CFRD, aminoglycoside therapy, age, and eGFR demonstrated a significant inverse relationship between uKIM-1 and %FEV1 that is independent of these potential confounders (Supplemental Table 5).

### Urinary neutrophil activation and lung disease exacerbation positively correlate with levels of renal injury markers

Neutrophils are highly abundant innate immune cells in the infected CF lung ^13,25^. In other models of local tissue injury neutrophils have been shown to disseminate to distal organs, including our prior findings that neutrophils migrate to the kidney following skin injury^15,26^. To ask if neutrophil activation is associated with kidney injury in PwCF, we quantified urine levels of calprotectin (S100A8/9) and neutrophil extracellular traps (NETs), known markers of neutrophil activation ^20^. Urinary calprotectin and/or NETs levels, normalized to creatinine, positively correlated (blue squares) with several urinary markers of tubular (NGAL, KIM-1, TFF3, β-NAG) and glomerular (PDX) injury (**Fig. 3A**). Levels of urinary calprotectin were positively correlated with total urine protein (**Fig. 3B**), further suggesting a link between neutrophil activation and renal injury. To understand the contribution of pulmonary exacerbation, which was present in 37.5% of PwCF at the time of urine collection, we performed PCA of urinary injury markers in relation to the pulmonary exacerbation (PEx) status. The urinary profile in 12/18 patients with a pulmonary exacerbation resembled that of patients without an exacerbation but there was a group of 6 patients with a pulmonary exacerbation that clustered separately (**Fig. 3C**). These 6 patients (encircled) had high levels of uNGAL, a tubular injury marker that is also secreted by activated neutrophils (**Fig. 3C-D**). Both urinary calprotectin and NGAL were increased in patients with an active PEx at the time of sample collection, as was total urine protein (**Fig. 3D**). Together, these observations support the model by which kidney injury in PwCF may be related to neutrophil activation at the time of lung exacerbation.

**Figure 3.**
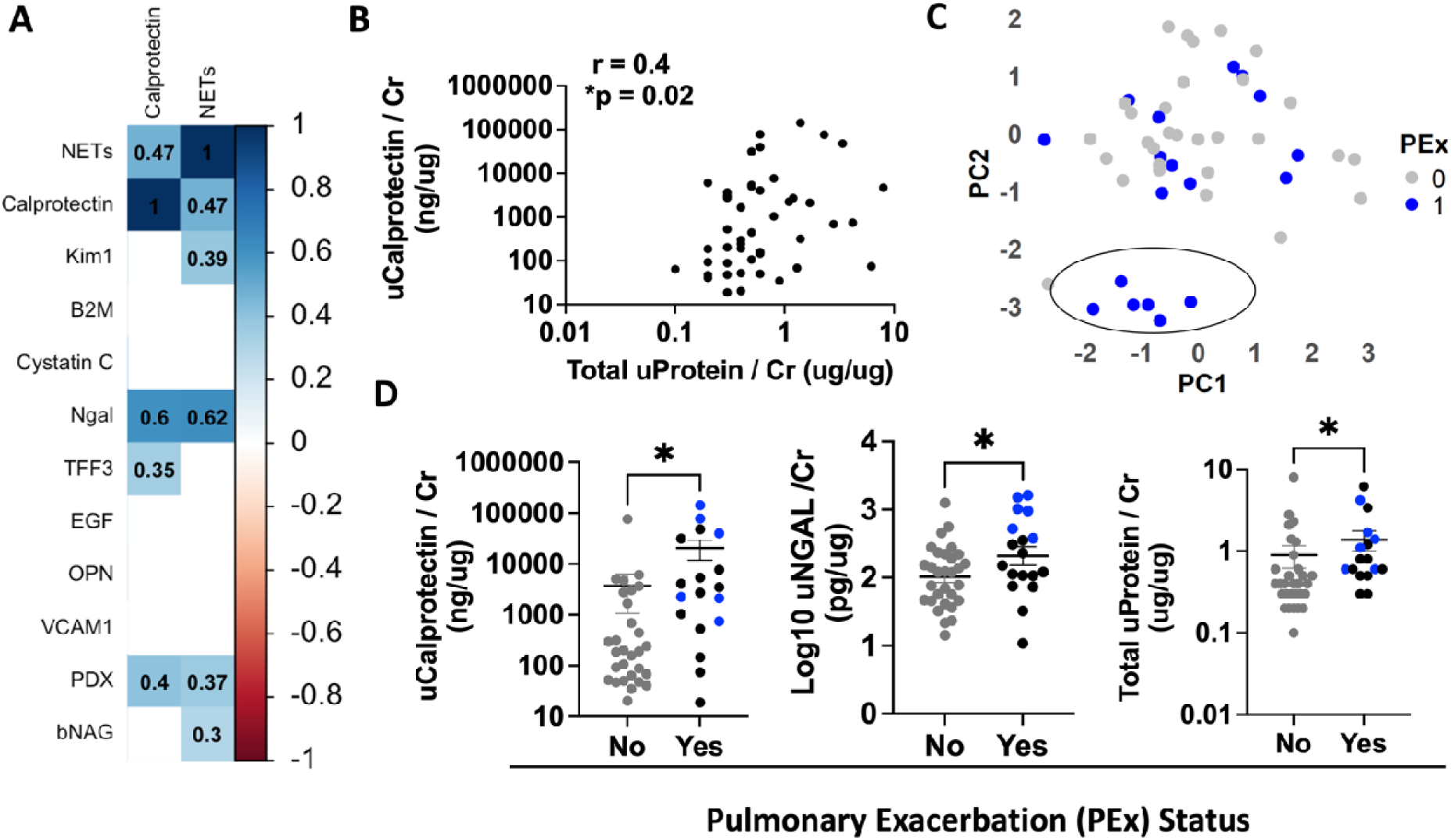
High urinary levels of tubular injury markers correlate with urinary calprotectin and NETs, in the context of pulmonary exacerbation. **(A)** Non-parametric Spearman correlation matrix of urinary kidney injury markers measured in Figure 1C, normalized to urine creatinine, with urinar markers of neutrophil activation: neutrophil extracellular traps (NETs, MPO: DNA complexes) and calprotectin (S100A8/A9), normalized to creatinine. The color of the squares indicates a significant positive (blue) or negative (red) correlation, with the r coefficient shown within each square (n = 48). **(B)** Non-parametric Spearman correlation analysis between urinary calprotectin (uCalprotectin) and total urine protein, both normalized to urine creatinine (Cr) (n = 48). **(C)** Robust principal component (PC) analysis of urinary markers measured in C in the CF cohort stratified by the status of the pulmonary exacerbation (PEx): present at the time of urine sample collection (blue) or no PEx at the time of sampl collection (grey). **(D)** Urinary levels of calprotectin, NGAL, and total protein, normalized to urine creatinine (Cr), in PwCF segregated by the state of pulmonary exacerbation at the time of urine sampl collection. Statistical significance determined by Student’s t-test or non-parametric t-test (n=48, *p < 0.05, **p < 0.01).

### Urinary neutrophil levels are elevated in CF patients with decreased lung function

While increased urinary calprotectin and NETs implicate neutrophil activation in renal injury in PwCF (**Fig. 3**), these are indirect measurements of neutrophil presence. Since recent studies have demonstrated that urine cell profiles closely mimic cellular diversity in the kidney tissue ^27,28^, we quantified neutrophil levels in the urine of PwCF by leveraging cell-specific DNA methylation (DNAm) signatures ^29^. As cells commit to different lineages, they inherit characteristic DNAm signatures in their epigenome (Supplemental Figure 1). Utilizing reference-based cell-type deconvolution of differentially methylated regions (DMRs), we quantified immune cells in the urine cell pellets from 11 PwCF (**Fig. 4A**). For these studies, we recruited a second cohort of PwCF as the samples collected in the initial CF cohort did not include urinary cell pellets (patient characteristics in Supplemental Table 3). Deconvolution of urinary DNAm data identified myeloid cells (Myel: monocytes and macrophages), neutrophils (Neu), memory B cells (Bmem), and regulatory T cells (Treg) as the most prevalent immune populations in the urine cell pellets of PwCF (**Fig. 4B**). Among innate immune cells, the percentage of neutrophils within all immune cells was significantly higher than that of myeloid cells (**Fig. 4C**). Interestingly, the percentage of neutrophils in urine immune cells inversely correlated with %FEV_1_ (**Fig. 4D**). No relationship was observed between myeloid cells, Treg, or memory B cells with %FEV_1_ (**Fig. 4D**). These findings suggest that increased neutrophil presence in the urine is associated with worse lung function.

**Figure 4.**
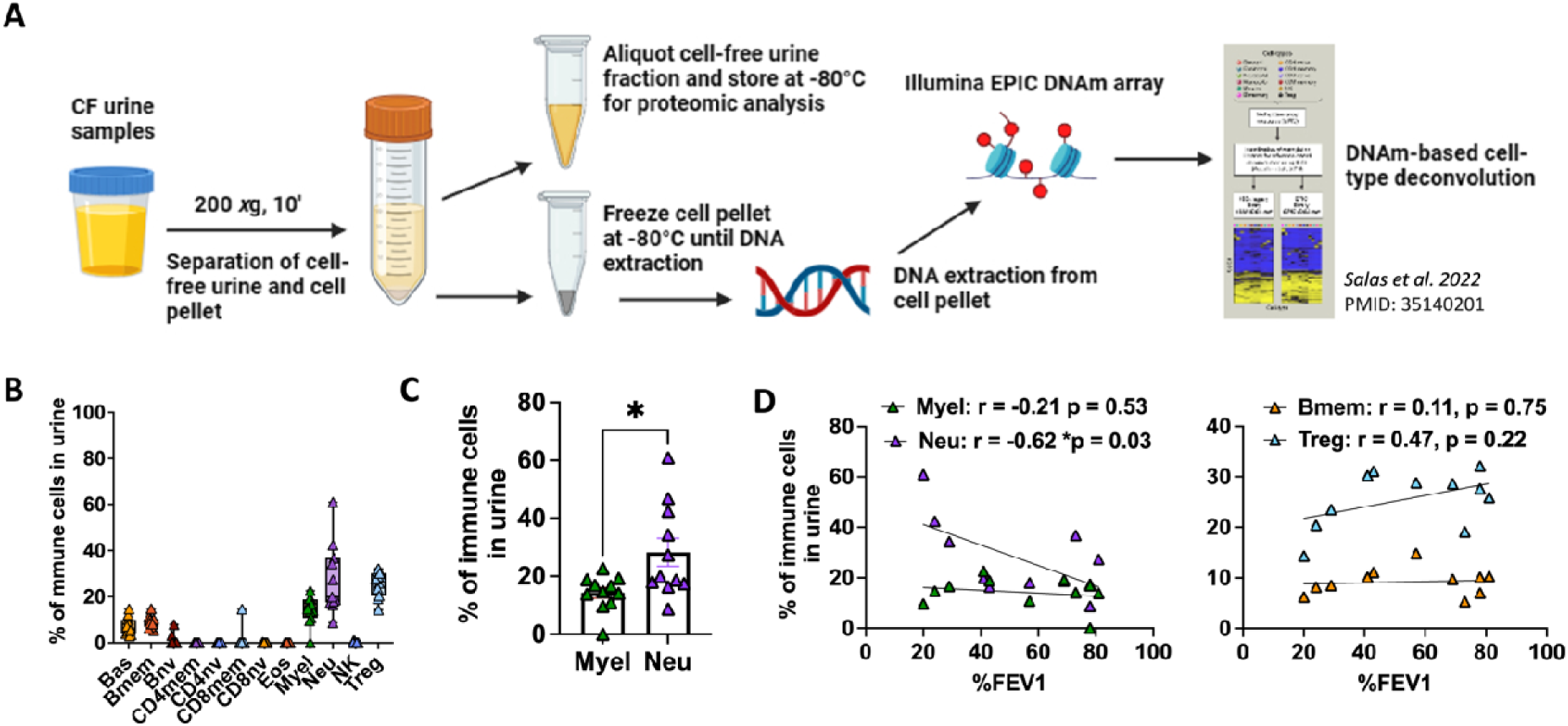
Urinary neutrophil levels correlate with worse lung function in PwCF. **(A)** Urine cell pellets and cell-free supernatants were separated by centrifugation. DNA was extracted from frozen cell pellets, and 250ng was run on an Illumina EPIC DNA methylation (DNAm) array for deconvolution of different immune cell populations. **(B)** Deconvolution of DNAm data demonstrated the presence of neutrophils (Neu), myeloid cells (Myel: monocytes and/or macrophages), regulatory T cells (Treg), memory B cells (Bmem), and basophils (Bas) in the urine of PwCF, within all immune cells. **(C)** The percentages of neutrophils and myeloid cells within all immune cells in urine were compared by Student’s t-test, n = 11, *p < 0.05. **(D)** Pearson’s correlation analyses between the percentage of Neu and Myel cell within urine immune cells (left) or the percentage of Treg and Bmem (right) and percent predicted forced expiratory volume in 1 second (%FEV1, n=11).

## Discussion

In this study, we report that PwCF display signs of renal injury, including increased levels of total urine protein in ∼29% and several urinary tubular injury markers (KIM-1, TFF3, β2MG, cystatin C, and β-NAG) in 17–48.9% of PwCF, in the absence of albuminuria. Lower urinary EGF correlated with decreased eGFR, suggestive of declining repair capacity of tubular epithelial cells, an indicator of progressive kidney injury. The increased urinary levels of most renal injury markers were independent of eGFR and remained increased in the CF cohort with normal eGFR (> 90). Though CFRD was not associated with higher urinary levels of any renal injury markers, there was a trend in higher total urine protein in CF patients with CFRD. Moreover, increased uVCAM-1 in the diabetic group could be due to endothelial cell dysfunction resulting from hyperglycemia. Only a subgroup of urinary injury markers (β2MG, cystatin C, and TFF3) were associated with antibiotic treatment. Together, these findings suggest that different factors likely play a role in urinary tubular injury and contribute to differential early urinary injury profiles: treatment, CFRD, kidney-intrinsic abnormalities due to the CFTR mutation, or potentially the response to disease. Our studies reveal several novel associations that link lung inflammation to renal injury in CF: i) increased uKIM-1 and neutrophil levels in urine immune cells inversely correlate with lung function; ii) urinary levels of kidney injury markers and proteinuria positively correlate with urinary markers of neutrophil activation, and iii) urinary levels of renal injury markers are elevated (KIM-1) or decreased (EGF) in PwCF with *P. aeruginosa* infection. Though neither aminoglycoside therapy nor CFRD were independently associated with increased uKIM-1 or decreased uEGF, future larger studies are required to define the contribution of CFRD simultaneous with aminoglycoside treatment to renal injury following a pulmonary exacerbation.

Aminoglycoside treatment has been associated with increases in numerous kidney injury markers acutely and their frequent administration could have chronic effects on renal injury. In our cohort, no aminoglycosides were administered intravenously and only three kidney injury markers (b2M, Cystatin C, and TFF3) were significantly correlated with tobramycin treatment, though this may be reflective of a small retrospective cohort. Since aminoglycosides are usually administered to treat lung infections and 38% of our cohort had a pulmonary exacerbation at the time of sample collection, lung inflammation could be a contributing factor to renal injury. Interestingly, increased urinary levels of these injury markers in PwCF correlated with higher levels of urinary calprotectin and NETs, markers of neutrophil activation, suggesting a relationship between inflammation and renal injury in a subset of PwCF. Neutrophils are known inflammatory responders and are abundant in the CF airway. In other settings of local injury, neutrophils have been shown to disseminate to other organs, including the kidney^15^. In our studies, uCalprotectin was associated with total urine protein levels, suggesting a relationship between neutrophil activation and renal injury. While it is difficult to ascertain whether the neutrophils are derived from infected and/or inflamed lungs, we found that %FEV_1_ is inversely correlated with the levels of neutrophils in urine cell pellets. Studies in ARDS and COPD demonstrate that inflammation in the lung can have both acute and chronic consequences on the kidney ^10,30^. However, if and by which mechanism neutrophils may mediate renal injury CF and whether subpopulations of PwCF would be more susceptible to neutrophil-mediated injury is unknown. Our analysis revealed a small population of CF patients with a pulmonary exacerbation who had high urinary levels of NGAL, a urine activation as well as a tubular injury marker. That uKIM-1 and uEGF levels were correlated with *P. aeruginosa* but not *S. aureus* infection may point to different contributions of lung pathogens to renal injury.

Another important contributor to the renal susceptibility to injury in PwCF may be aberrant CFTR function in the kidney. In addition to the apical surfaces of proximal and distal tubules, CFTR is also highly expressed in the apical endosomes, where it plays a role in the endocytic uptake of low-molecular-weight (LMW) proteins ^31^. The defect in endocytosis of LMW proteins by the proximal tubular cells in the CFTR-deficient mice is attributed to decreased levels of cubilin ^31^. Therefore, the accumulation of proteins such as β2MG, TFF3, or NGAL, in the urine of PwCF may in part be a consequence of intrinsic renal CFTR defects. Studies of lung epithelial and endothelial cells have demonstrated increased expression of adhesion molecules, neutrophil chemoattractants (IL-8, IL-1b), and inflammatory cytokines in the absence of a functional CFTR ^32^. Moreover, high peribronchial neutrophil infiltration in CF infants prior to any infection ^33,34^, suggests that renal endothelial and epithelial intrinsic defects in the CFTR may also contribute to neutrophil recruitment and activation in the kidney. We were not able to capture any CFTR mutation-specific effects due to the small number of non-Class II PwCF and the small number of patients treated with the triple CFTR modulator in this retrospective analysis. However, even in a CF cohort predominantly treated with the triple CFTR modulator (n = 9/11), we detected a wide range of neutrophil levels in urine, which was strongly linked with lung function (Fig. 4). These data suggest that inflammation and neutrophil infiltration, linked to worsened lung function and pulmonary exacerbation (36% of this cohort had a pulmonary exacerbation), may still be contributing to renal injury even in patients treated with HEMT.

The limitations of this study include the relatively small sample size and the cross-sectional design, which limit our ability to establish causality and the contribution of each clinical parameter to the observed trends. The lack of associations between some urinary markers with CFRD or aminoglycoside treatment could be due to the small sample size. Studies of larger cohorts will further delineate the impact of CFRD, treatment due to acute illness, the cumulative impact of prior aminoglycoside therapy on kidney injury in CF, and potential additive effects of aminoglycoside therapy and CFRD. Urine samples in CF cohort 1 and matched healthy controls were directly frozen and thus some of the proteins could be derived from cells in urine. Since leukocytes, besides renal tubular epithelial cells, can also produce KIM-1, and activated neutrophils can make NGAL/Lipocalin-2, future studies will define the cellular origin of these markers via analyses of urinary exosomes. Moreover, future studies with larger cohorts and longitudinal design are necessary to better elucidate the effect of treatment type and duration, lung disease flares, and co-morbidities on the underlying kidney injury in PwCF. While the methylation analysis of urine represents a non-invasive way to ask how immune cells may contribute to renal injury signatures, it is plausible that the same methylation signature could identify neutrophils of different functionalities (e.g. low-density immature, normal-density mature, and MDSCs). Thus, future studies will implement more specific cellular methylation profiles of urinary neutrophil subsets. Due to the size of the cohort, sex as a variable was not carefully considered and will be integrated into statistical analyses in future studies. Future studies will investigate the effects of HEMT on renal injury processes.

## Data Availability

All data produced in the present study are available upon reasonable request to the authors

## Acknowledgments

Luminex urine analyses were carried out in DartLab, the Immune Monitoring and Flow Cytometry Shared Resource at the Norris Cotton Cancer Center at Dartmouth, with NCI Cancer Center Support Grant 5P30 CA023108-41. DNA methylation array study was carried out in the Genomics and Molecular Biology Shared Resource (RRID:SCR_021293) at Dartmouth which is supported by NCI Cancer Center Support Grant 5P30CA023108 and NIH S10 (1S10OD030242) awards. This work was supported by National Institutes of Health (5 P20GM130454-04), Dartmouth Cystic Fibrosis Pilot Program (P30DK117469), and Cystic Fibrosis Foundation (SKOPEL24A0-MPI) to S.S.G. and National Institutes of Health (R01 HL151385 and P30 DK117469), Cystic Fibrosis Foundation (STANTO19R0), and the Flatley Foundation to B.A.S.

## Supplemental Material

**Supplemental Table 1:**
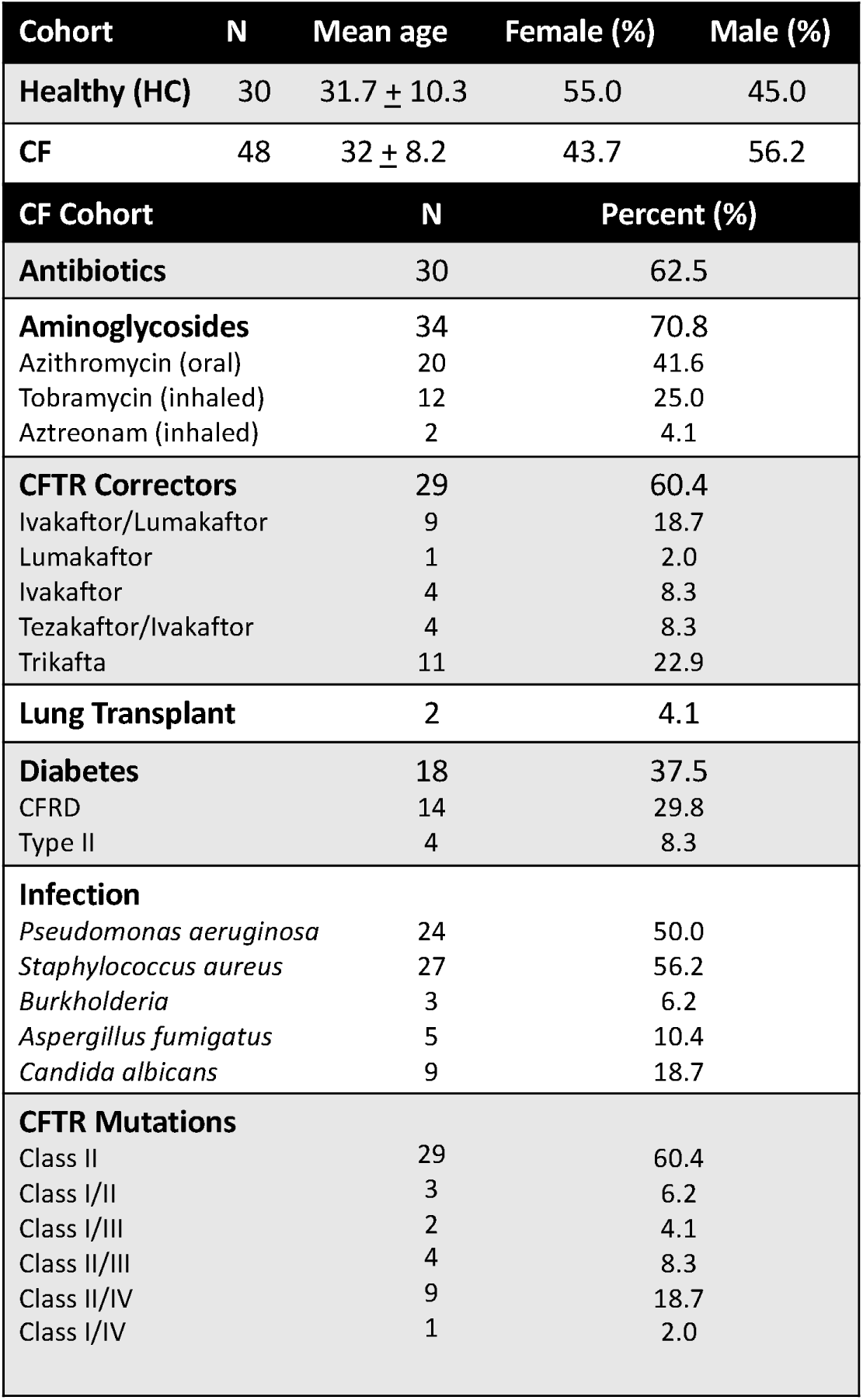
Characteristics of the CF and Healthy Cohorts.

**Supplemental Table 2:** Clinical parameters of the CF cohort.

| Clinical covariates | Range in CF cohort #1<br>(Average ± SD) | Normal range |
| --- | --- | --- |
| eGFR | 106.4 ± 66.34 | ≥ 90 mL/min/1.73m<sup>2</sup> |
| %FEV1 | 71.8 ± 22.6 | >80% |
| *Pulmonary Exacerbation* | 37.5% |  |
| Hb1Ac | 6.7 ± 2.1 | 4–5.6% |
| Hematocrit | 42.5 ± 2.57 | Male: 38.3–48.6% |
| | 37.7 ± 3.92 | Female: 35.5–44.9% |
| Serum bicarbonate | 25.8 ± 3.54 | 22–29 mEq/L |

**Supplemental Table 3:** CF Cohort 2 (Figure 4) patient characteristics.

| CF Cohort | N | Percent (%) |
| --- | --- | --- |
| **Antibiotics** | 5 | 45.4 |
| Azithromycin | 4 | 36.3 |
| Tobramycin | 3 | 27.2 |
| Aztreonam | 4 | 36.3 |
| **CFTR Correctors** | 9 | 81.8 |
| Eleacaftor/tezacaftor/ivacaftor | 9 | 81.8 |
| **CFRD** | 7 | 63.6 |
| **Infection** | 6 | 54.5 |
| *Pseudomonas aeruginosa* | 3 | 27.2 |
| *Staphylococcus aureus* | 3 | 27.2 |
| **CFTR Mutations** |  |  |
| Class II | 6 | 54.5 |
| Class II/III | 3 | 27.2 |
| Class II/IV | 2 | 18.1 |
| Clinical covariates | Range in CF cohort #2<br>(Average ± SD) | Normal range |
| eGFR | 116.63 ± 24.73 | ≥ 90 mL/min/1.73m<sup>2</sup> |
| %FEV1 | 54 ± 23.34 | >80% |
| *Pulmonary Exacerbation* | 36.4% |  |
| Hb1Ac | 6.7 ± 2.1 | 4–5.6% |
| Hematocrit | 44 ± 0.89 | Male: 38.3–48.6% |
| | 37.93 ± 7.20 | Female: 35.5–44.9% |
| Serum bicarbonate | 25.9 ± 2.64 | 22–29 mEq/L |

**Supplemental Table 4:** Correlation analysis of urinary kidney injury markers and clinical parameters in PwCF.

| CF N=48 | Total Urine Protein | uβ2M | uCystatin C | uNGAL | uKim-1 | uTFF3 | uβ-NAG | uVCAM1 | uEGF | uOPN | uPDX |
| --- | --- | --- | --- | --- | --- | --- | --- | --- | --- | --- | --- |
| eGFR | - | - | - | - | - | - | - | - | 0.02* | - | 0.001* |
| %FEV1 | - | - | - | - | 0.01* | - | - | - | - | - | - |
| Hypertension | - | - | - | - | - | - | - | 0.02* | 0.0004*** | 0.04* | - |
| Diabetes | - | - | - | - | - | - | - | 0.02* | - | - | - |
| HbA1c | - | - | - | - | - | - | - | 0.02* | - | - | - |
| Hematocrit<sup>#</sup> | 0.007** | - | 0.01* | 0.002** | - | - | - | - | - | 0.01* | 0.0005*** |
| Serum Bicarb | - | - | - | - | - | - | - | - | - | - | - |
| Lung transplant | - | - | - | - | - | - | - | - | - | - | - |
| BMI | - | - | - | - | - | - | - | - | - | - | - |
| Age | - | - | - | - | - | - | - | 0.03* | 0.0005*** | - | - |
| Sex |  |  |  | <0.001*** |  |  |  |  | 0.01* |  | <0.0001* |
| Azithromycin | - | - | - | - | - | 0.03* | - | - | - | - | - |
| Tobramycin |  | 0.03* | 0.005** | - | - | 0.0008*** | - | - | - | - | - |
| Aztreonam | - | - | - | - | - | 0.04* | - | - | - | - | - |
| Elezaftor/<br>tezacaftor/<br>ivaftor | - | - | - | - | - | - | - | - | - | 0.01* | - |

<sup>#</sup> All correlations with hematocrit levels are negative
β2m = β2-microglobulin, Kim-1 = kidney injury marker 1, EGF = epidermal growth factor, OPN = osteopontin, NGAL = Lipocalin, TFF3 = Trefoil factor 3, PDX = Podocalyxin, βNAG = N-acetyl-β-d-glucosaminidase; BMI = Body Mass Index, HbA1c = Hemoglobin A1c;
All urine measurements normalized to urine creatinine

**Supplemental Table 5:** Multivariable regression analysis of uKIM-1 and %FEV1 adjusted for CFRD, Aminoglycosides (Tobi), Age, and eGFR.

| Parameter estimates | Variable | Estimate | Standard error | 95% CI (asymptotic) | \|t\| | P value | P value summary |
| --- | --- | --- | --- | --- | --- | --- | --- |
| β0 | Intercept | 1.190 | 0.3670 | 0.4494 to 1.931 | 3.243 | 0.0023 | ** |
| β1 | % FEV1 | -0.004947 | 0.002155 | -0.009296 to -0.0005979 | 2.296 | 0.0268 | * |
| β2 | CFRD[0.0] | -0.05190 | 0.1004 | -0.2545 to 0.1508 | 0.5168 | 0.6080 | ns |
| β3 | Tobi[1] | 0.01490 | 0.1089 | -0.2048 to 0.2346 | 0.1369 | 0.8917 | ns |
| β4 | Age | -0.001697 | 0.006249 | -0.01431 to 0.01091 | 0.2715 | 0.7873 | ns |
| β5 | eGFR | -0.0001061 | 0.001524 | -0.003181 to 0.002969 | 0.06963 | 0.9448 | ns |

### Supplemental Figures

**Supplemental Figure 1:**
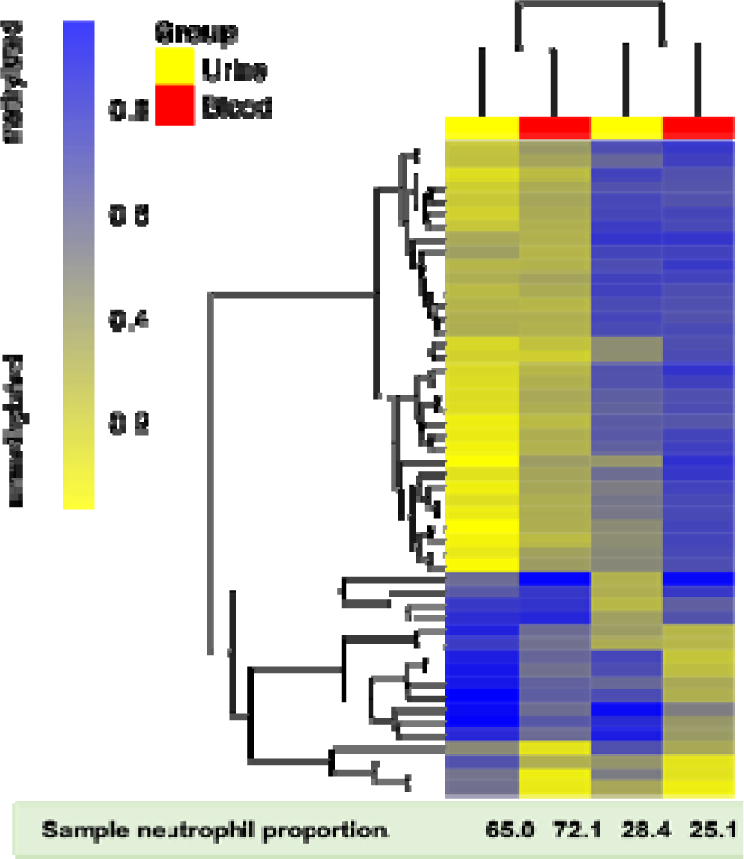
Unbiassed clustering of blood and urine samples using 50 CpG sites with known neutrophil-specific DNAm status results in grouping by neutrophil proportion, not specimen type.

**Supplemental Figure 2:**
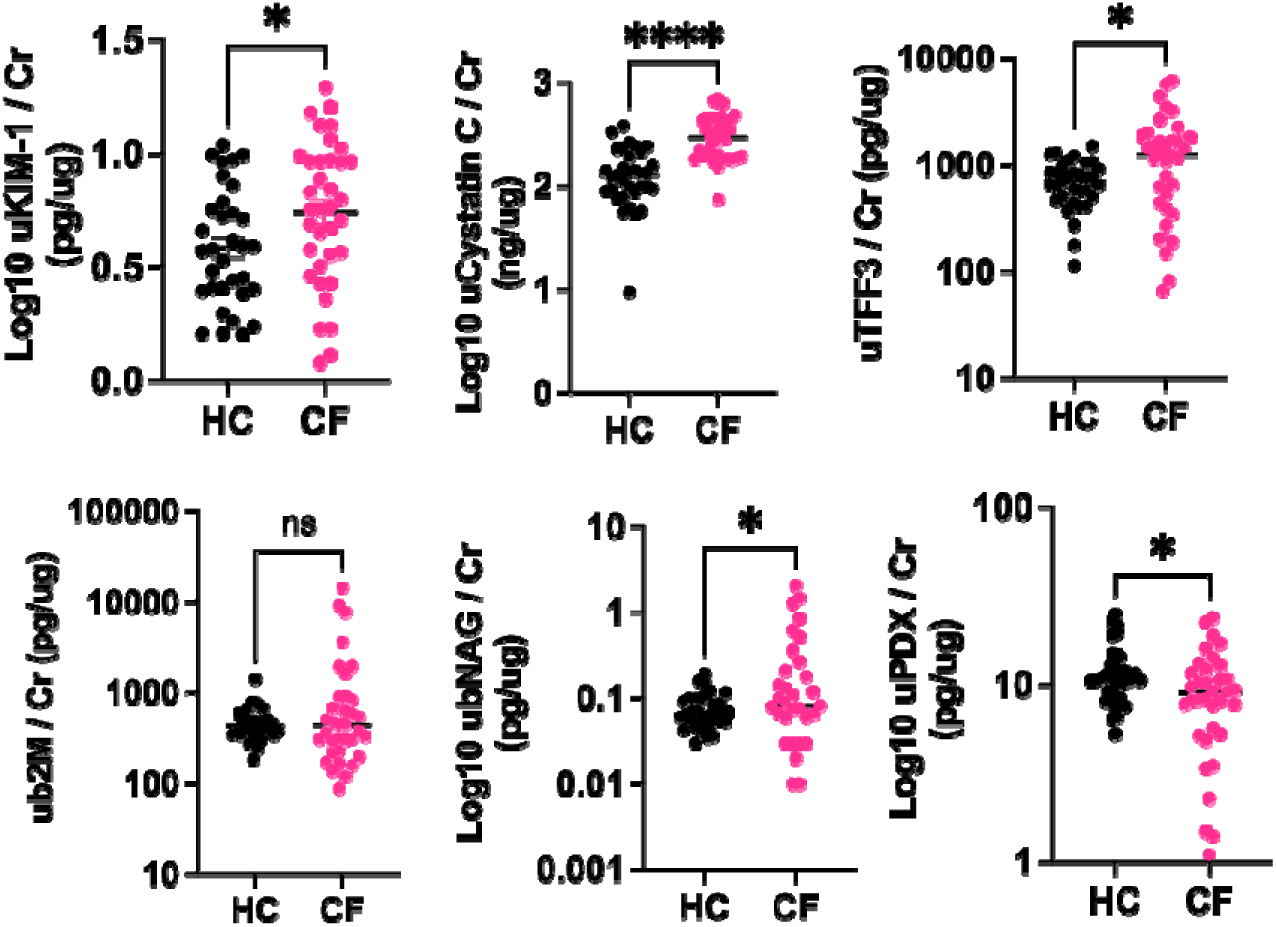
Concentration of urinary kidney injury markers in healthy controls (HC, n = 33) and PwCF with eGFR > 90 (CF, n = 35). normalized to urine creatinine. Significance was determined non-parametric t-test for not normally distributed data or Student’s t-test for lognormally and normally distributed data: *p<0.05, ****p < 0.0001.

**Supplemental Figure 3:**
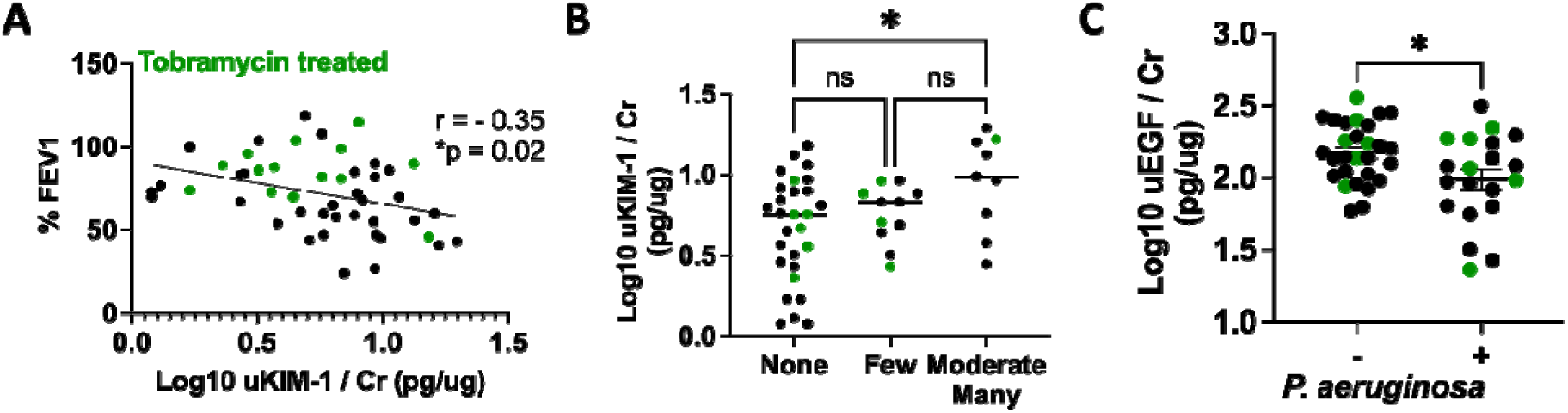
Treatment with Tobramycin (green dots) at the time of sample collection is not associated with higher uKIM-1 (A-B) or lower uEGF (C) in patients infected with P. aeruginosa. uKIM-1 = Urinary Kidney Injury Marker 1; uEGF = urinary Epidermal Growth Factor. Statistical significance defined by (A) Pearson’s correlation analysis, (B) One-way ANOVA with Bonferroni post-hoc, and (C) Student’s t-test. *p < 0.05.

